# Eq-DIAGNOBATCH, a Shiny application for topical product equivalence with batch effect and multivariate outlier analysis

**DOI:** 10.1101/2024.12.16.24319103

**Authors:** Toni Monleón-Getino, Jordi Ocaña

## Abstract

The Eq-DIAGNOBATCH application presented here complements a previous theoretical article about establishing the equivalence of topical products in a parallel linear model, accounting for a batch effect or not. This application provides a very simple way of carrying out the equivalence test for this type of situation, efficiently obtaining the estimates of the parameters of interest (equivalence intervals, geometric mean ratio, and variance components), and allowing calculations to be performed assuming a batch effect or not. It also examines the statistical implications of the presence of outliers in the sample, from both a univariate and multivariate point of view. The operation and mode of use of Eq-DIAGNOBATCH are presented, including examples related with the quality and equivalence of topical products and bioequivalence analysis. Some of these examples come from the literature and others from real-life industry case studies.

## 1. Introduction

Eq-DIAGNOBATCH (Figure 1) is an application developed in R language (R Core Team, 2022) and encapsulated in a server web application using Shiny (Chang et al., 2022). The application was designed to complement the equivalence method developed by Ocaña et al. (2020) for equivalence in a two-factor design (fixed factor “form” and random factor “batch”). It has been extended with the possibility of analyzing a one-factor design (“form”) and detecting outliers in the datasets, using multivariate and univariate methods (Monleon-Getino et al, 2020; Monleón-Getino and Cavalleri, 2022). The main objective of this application is to promote the use of statistical methods and facilitate calculations to determine the statistical equivalence between a test product (T) and a reference product (R). The application is free to use and can be accessed at the following link: https://amonleongbiost3.shinyapps.io/EqDIAGNOBATCH/. The different usage dataframes can be found in https://github.com/amonleong/EqDIAGNOBATCH.

**Figure 1:**
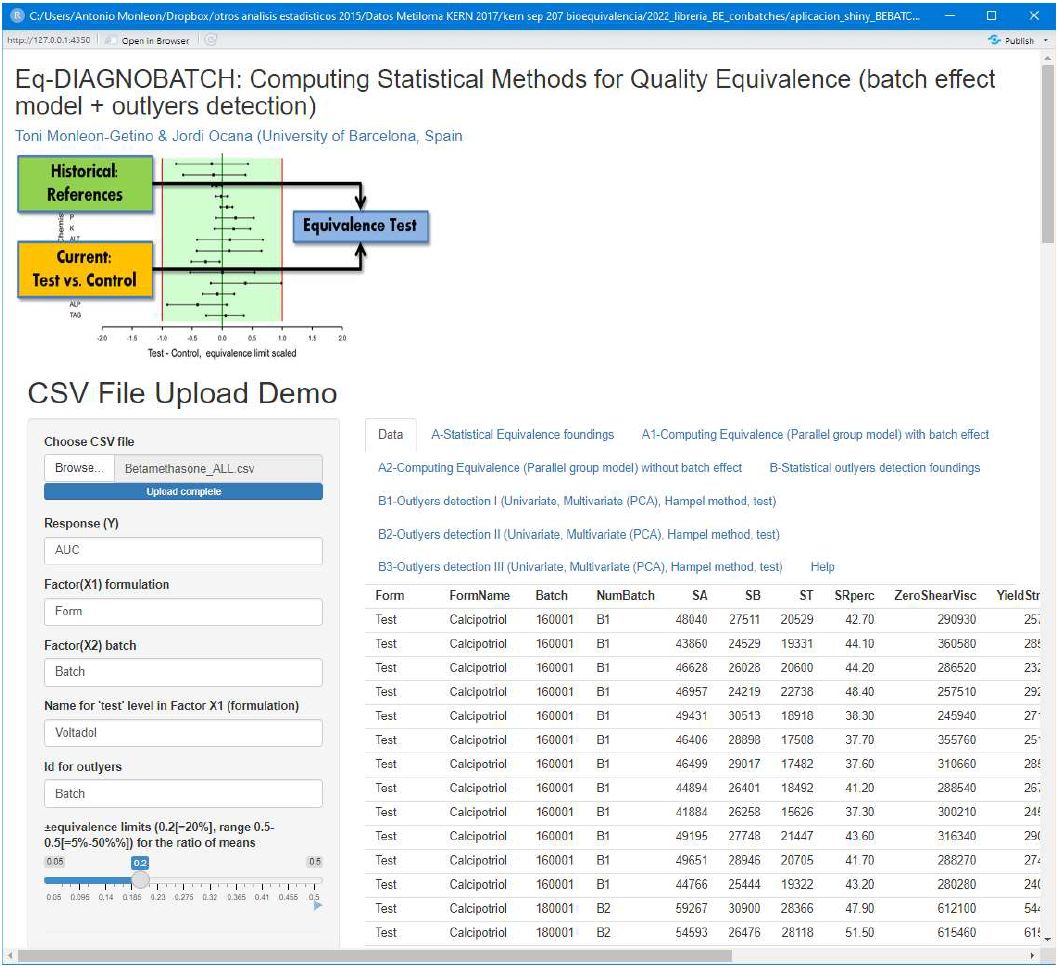
Central Screen of Eq-DIAGNOBATCH APP

Figure 1 shows the starting screen of the application, where the user loads a data matrix with a specific format related to the equivalence analysis. It is then necessary to provide the values of the arguments, specifying the process type (equivalence or outliers). The application automatically controls if they are in an appropriate format, as follows: a) pharmaceutical formulas (T,R)^1^; b) if there is a batch factor and the column name specifying it; c) the name of level T in the formulation column (paragraph a); d) if outliers are identified, the column name for groups (e.g., batches or identifiers), and finally, e) equivalence limits (0.2[=20%] within a range of 0.05-0.5[=5%-50%%]) for the ratio of means. For a detailed description of the equivalence test between product formulations, see Ocaña et al. (2020).

Figure 2 shows a flowchart of the different statistical processes performed by the application. It can be seen that Eq-DIAGNOBATCH is separated into two procedures: A) related to equivalence in the case of parallel groups, according to the model with a treatment factor and batch effect (A1) or without the batch effect (A2), and B) corresponding to the analysis of outliers in a multivariate (different response variables, PCA) and univariate case, through different exploratory graphs (Hampel method) and hypothesis test (Grubbs test).

**Figure 2:**
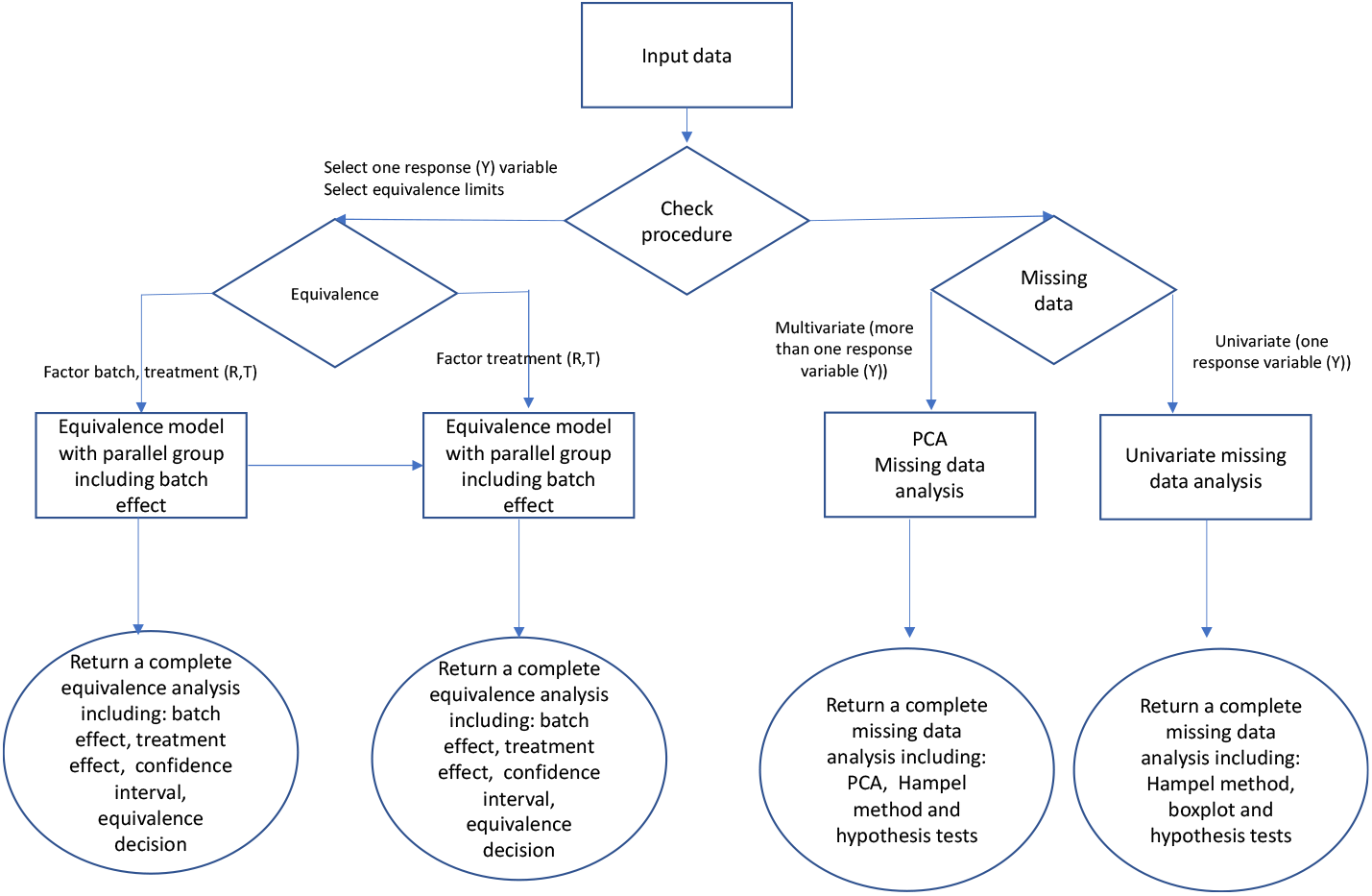
Flow chart of the Eq-DIAGNOBATCH application, with the two procedures: equivalence and outlier data analysis.

Figure 2 shows the execution flow of Eq-DIAGNOBATCH and how it is used in different procedures.

## 2. Statistical methods used in the application

Eq-DIAGNOBACTH is based on the theory described in Ocaña et al. (2020) and other texts about statistical tests of equivalence. This paper introduces an equivalence method based on a confidence interval for the ratio of the geometric means, under a linear model of parallel groups and a random batch effect (Annex A). Since sometimes data do not include multiple batches (or researchers prefer to ignore them), the models under consideration were complemented with a parallel group model without a batch effect (Annex A).

Outliers may occur in these datasets. The application also includes a collection of different outlier detection methods in data analysis (Hampel method and Grubbs test). Outlier detection can be performed using a univariate or multivariate approach (Annex B).

## 3. Usage examples

Some cases of usage can be found in https://github.com/amonleong/EqDIAGNOBATCH. They are based on the data files (csv format):

- Betamethasone_ALL.csv
- Betamethasone_ALL_PCA.csv:
- Betamethasone_REFERENCE.csv
- Betamethasone_TEST.csv
- Eq_Parallel_withoutBatch.csv
- Eq_Parallel_Batch.csv
- Analysis_outliers_univ.csv
- Analysis_outliers_multiv.csv

Betamethasone_ALL.csv contains data from all response variables used by Ocaña et al (2020). Betamethasone_ALL_PCA.csv contains PCA1 and PCA2 as response variables obtained by principal components of all response variables from Betamethasone_ALL.csv. The file Betamethasone_REFERENCE.csv contains only the reference group (R) and Betamethasone_TEST.csv contains only the test group (T). These first four files can be used both for equivalence analysis testing (with and without a batch effect), as well as for multivariate outlier analysis (provided they contain more than one response variable). The Eq_Parallel_withoutBatch2.csv file contains data from a parallel group equivalence experiment that includes a batch effect with a single response variable (Var). Eq_Parallel_withoutBatch.csv contains data from a parallel group equivalence experiment that does not include a batch effect with a single response variable (area under the curve, AUC). Analysis_outliers_univ.csv contains data from different batches and a single response variable (var3) for performing univariate outlier analyses. Finally, Analysis_outliers_multiv.csv contains data from different batches and three response variables (var1, var2, var3) for performing multivariate outlier analysis.

Some examples of analyses carried out using Eq-DIAGNOBATCH are discussed below.

### 3.1. Establishing equivalence for parallel groups with a batch effect

In this case, the file Eq_Parallel_Batch.csv will be used with data from a topical product with the aim of establishing equivalence through an experiment in which a parallel group model with a batch effect (random) is generated. Eight different batches can be observed (batch column in Figure 2). The calculations are based on the equivalence for the ratio of means, and the equivalence limits are specified as an argument (see Figure 2). The clinical variable to establish equivalence is referred to here as VAR, but in practice it can be AUC or similar.

Figure 2 shows the data loaded in the application.

Figure 3 shows the main panel of the App Eq-DIAGNOBATCH and the arguments that need to be loaded are displayed on the left of the screen (Ex equivalence limits for the ratio of means).

**Figure 3:**
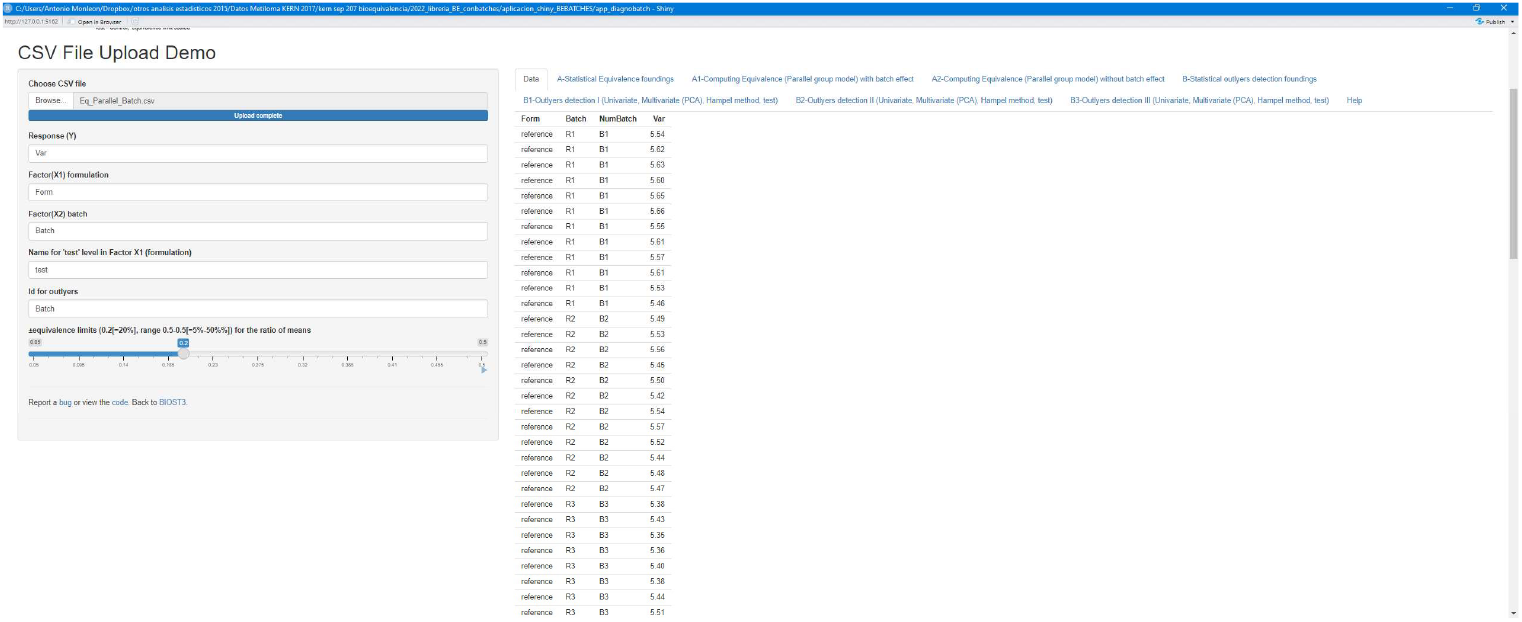
Eq-DIAGNOBATCH: Data from Eq_Parallel_Batch.csv uploaded in the application. On the left are the principal arguments for the calculations and different options (Tabs A for equivalence and B for outlier analysis).

The equivalence results are provided in a list at the bottom of the screen (Figs. 4 and 5) according to the equivalence model used (with or without a batch effect; Figures 4 and 5, respectively). The calculations are based on the equivalence for the ratio of means and the equivalence limits need to be specified as an argument (see Figure 2).

**Figure 4:**
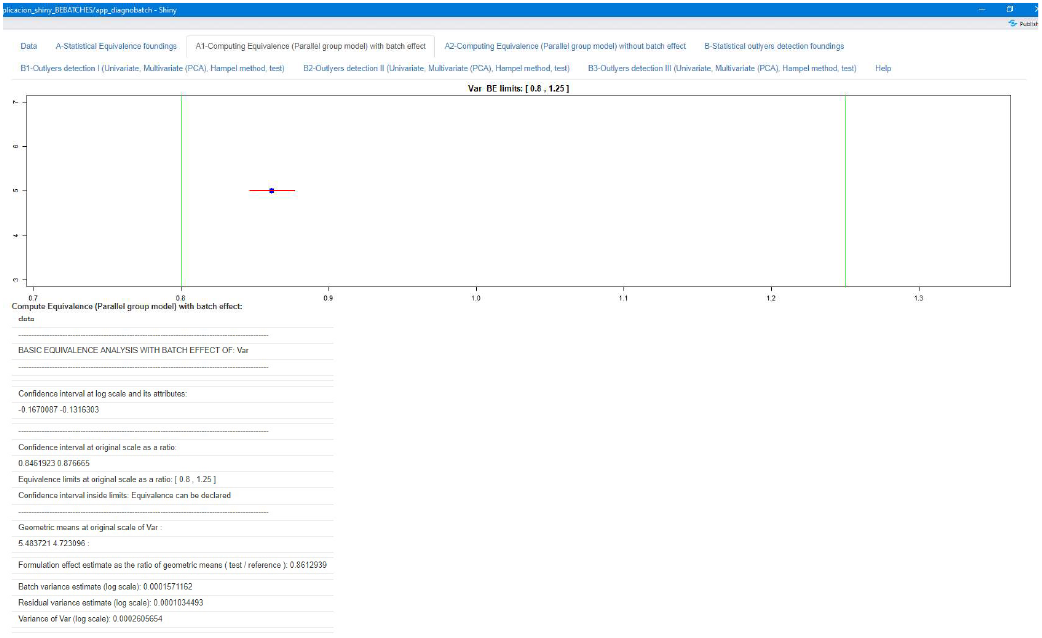
Eq-DIAGNOBATCH: Equivalence calculation for a parallel group design with a batch effect using equivalence intervals. Ratio of means estimation (blue dot), the equivalence limits (green lines) and equivalence confidence limits estimation (red line) are represented. Additionally, the statistical results of a complete equivalence test are shown.

**Figure 5:**
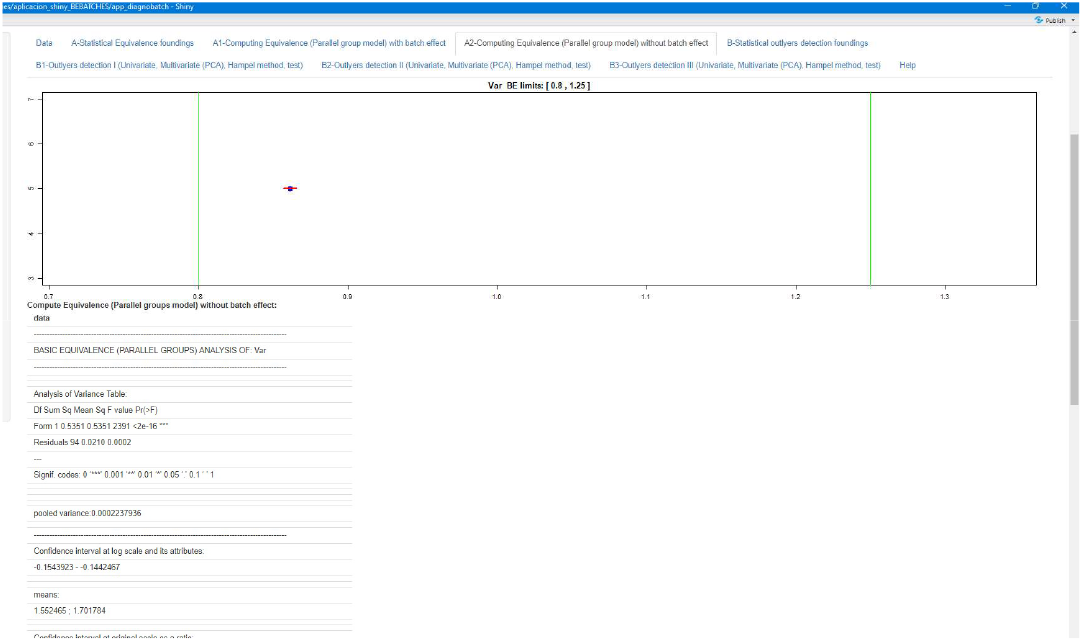
Eq-DIAGNOBATCH: Equivalence calculation for a parallel group design without a batch effect using equivalence intervals. Ratio of means estimation (blue dot), equivalence limits (green lines), and equivalence confidence limits estimation (red line) are represented. Additionally, the statistical results of a complete equivalence test are shown.

Regarding the list for the model with a batch effect, the following results can be seen on the application screen:

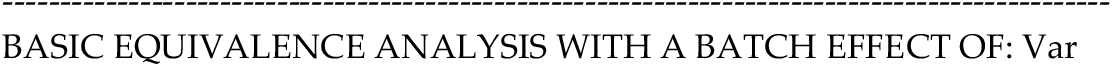
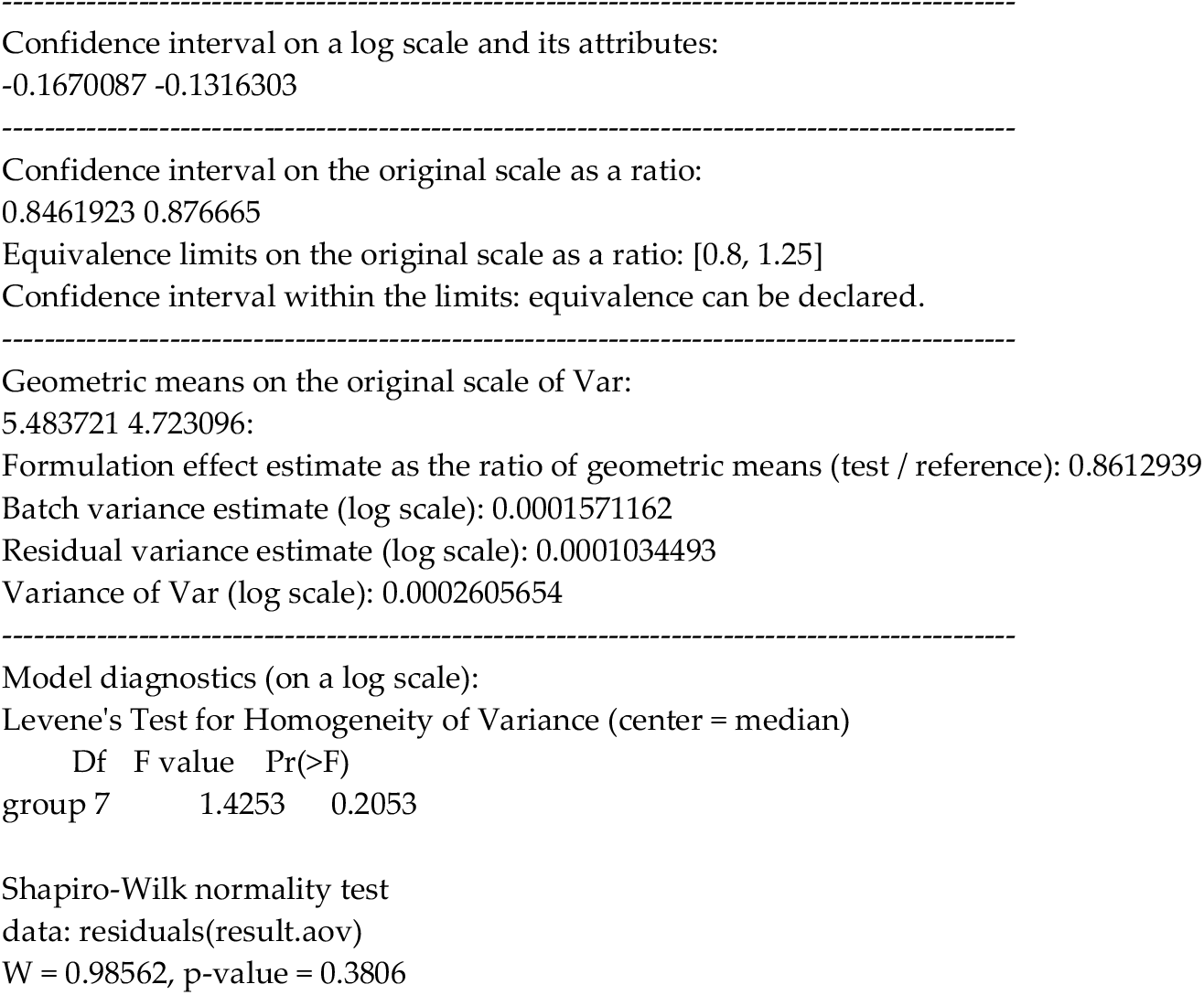

In this this case, equivalence can be declared because the confidence interval on the original scale for the ratio of geometric means is [0.8461923 0.876665] and it is fully within the equivalence limits on the original scale as a ratio: [0.8, 1.25]. The estimation of the formulation effect as the ratio of geometric means (test / reference) is 0.8612939. In addition, the calculations indicate there are no detectable problems with the homogeneity of variances (p=0.2053, Levene’s Test for Homogeneity of Variance) between batches (n=8 batches) and with the normality of data (p=0.3806, Shapiro-Wilk normality test). Finally, the procedure also computes the estimation of variance components for Batch variance and Residual variance on a log scale. The sum of variance components is the response variance (Var in this example).

### 3.2. Establishing equivalence for parallel groups without a batch effect

Now we are going to consider the same case but without a batch effect, using the previous example (using file Eq_Parallel_Batch.csv, response variable Var), i.e., only the levels of the treatment factor are compared. The observed results computed by the application are the following:

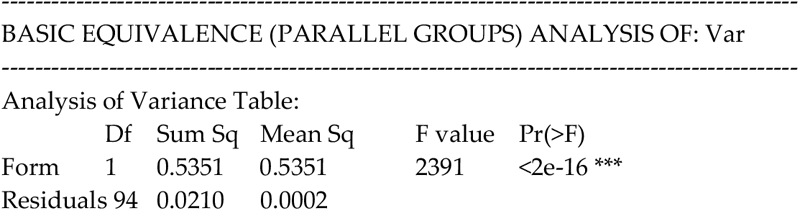
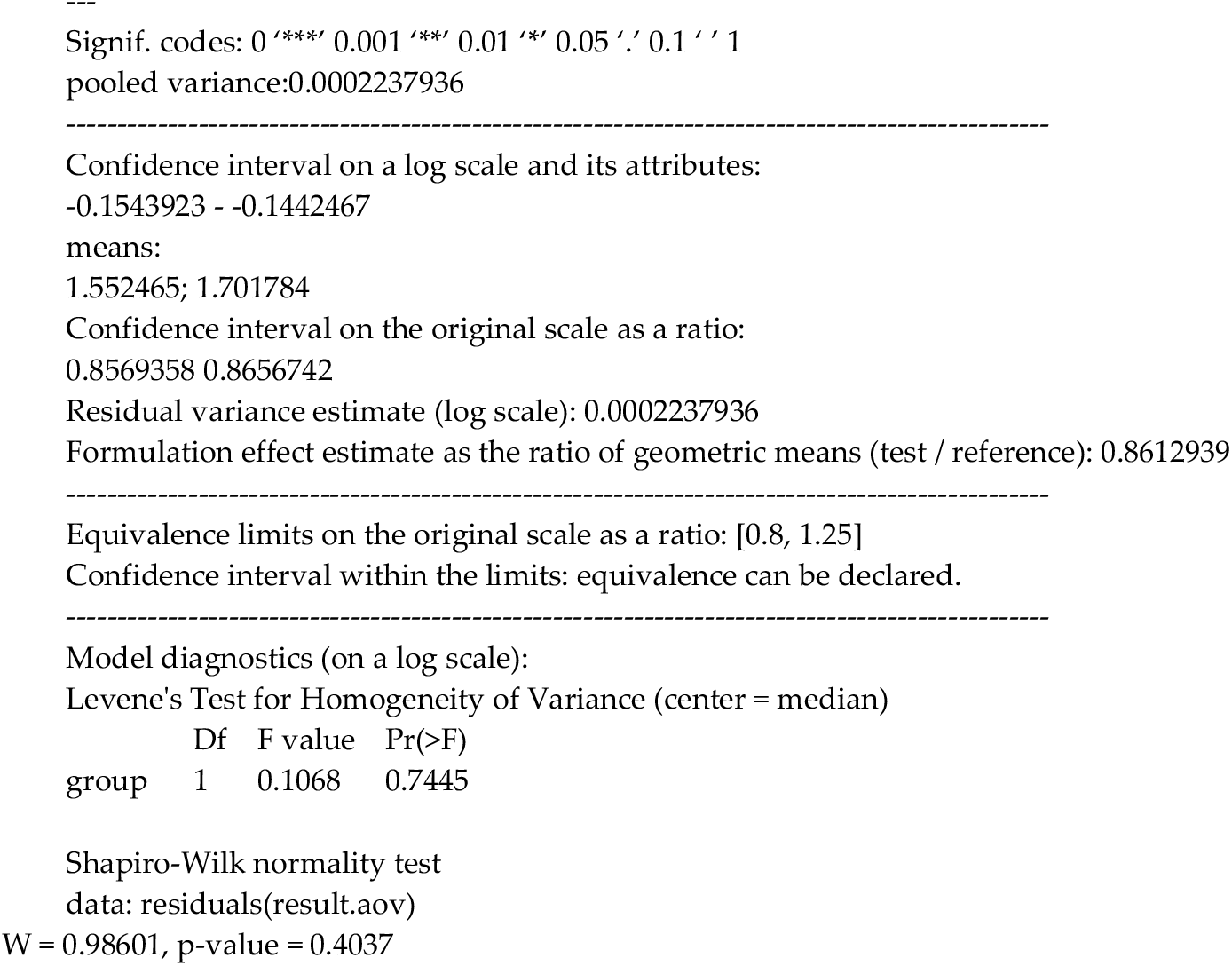

Equivalence can also be declared for this case because the confidence interval on the original scale for the ratio is [0.8569358 0.8656742], fully within the equivalence limits [0.8, 1.25]. The estimation of the formulation effect as the ratio of geometric means (test / reference) is 0.8612939. Again, the calculations indicate that there are no problems with the homogeneity of variance (p=0.7445, Levene’s Test for Homogeneity of Variance) between groups T/R (n=2) and with the normality of data analyzed (p=0.4037, Shapiro-Wilk normality test). Finally, the procedure also computes the estimation of variance components for Residual variance on a log scale.

Finally, we consider a case where it is not possible to declare equivalence between T and R in a parallel group design and without a batch factor, as in the file Eq_Parallel_withoutBatch.csv. First, the file must be loaded in the application and the equivalence limit specified. Next, the requested parameters must be specified, including the name of the variable of interest (AUC), the type T treatment, and the column that contains the treatment factor (Treatment). Then, the analysis is carried out and the results corresponding to the equivalence analysis without a batch effect can be observed in the tab (Figure 6).

**Figure 6.**
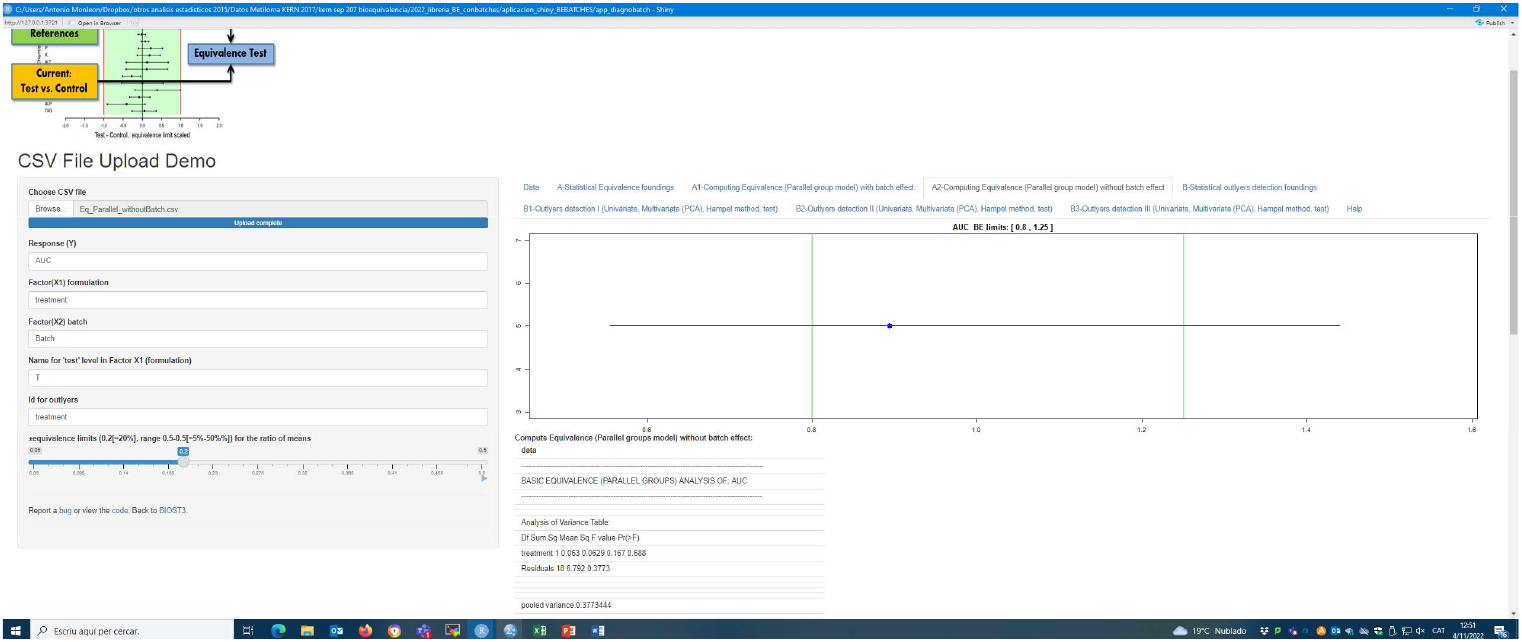
Eq-DIAGNOBATCH: Equivalence calculation in a parallel group design without a batch effect using equivalence intervals (file Eq_Parallel_withoutBatch.csv). Ratio of means estimation (blue dot), the equivalence limits (green lines), and equivalence confidence limits estimation (red line) are represented. Additionally, the statistical results of a complete equivalence test are shown.

Figure 6 shows the results of the equivalence analysis of the file Eq_Parallel_withoutBatch.csv. The data are from an equivalence experiment between two treatments (R / T) with a single response variable, where there are no batches. In the experiment, a huge variability of data was detected. It can be verified that equivalence cannot be established using the parallel group model (Figure 6), as the estimated confidence intervals are not fully within the interval [0.8, 1.25].

### 3.2. Univariate outlier analysis

Eq-DIAGNOBATCH also allows the analysis of outliers in a univariate or multivariate way (tabs named B in the outlier section in the application). The purpose of this is to find explanations for the non-declaration of equivalence as, on some occasions, highly variable batches and observations are found. These should be reviewed by the technical staff in case they need to be excluded from the equivalence analysis.

Here we present an analysis of outlier values carried out by the application for the same example as in the previous section, Eq_Parallel_withoutBatch.csv (where it was not possible to declare equivalence in the model without batches) with the aim of identifying samples that can cause statistical problems in our equivalence analysis. The analysis is based on the explanations provided in the introduction using methods commonly used by statisticians.

The results were:

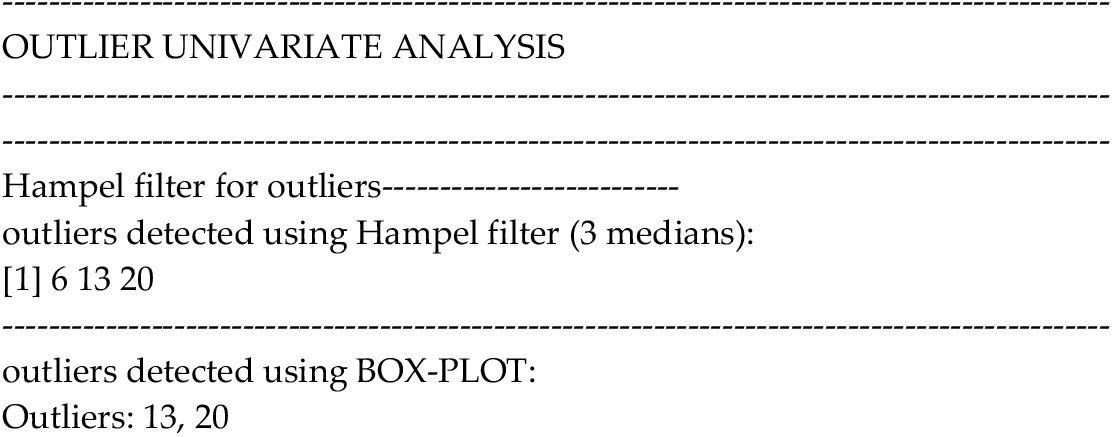
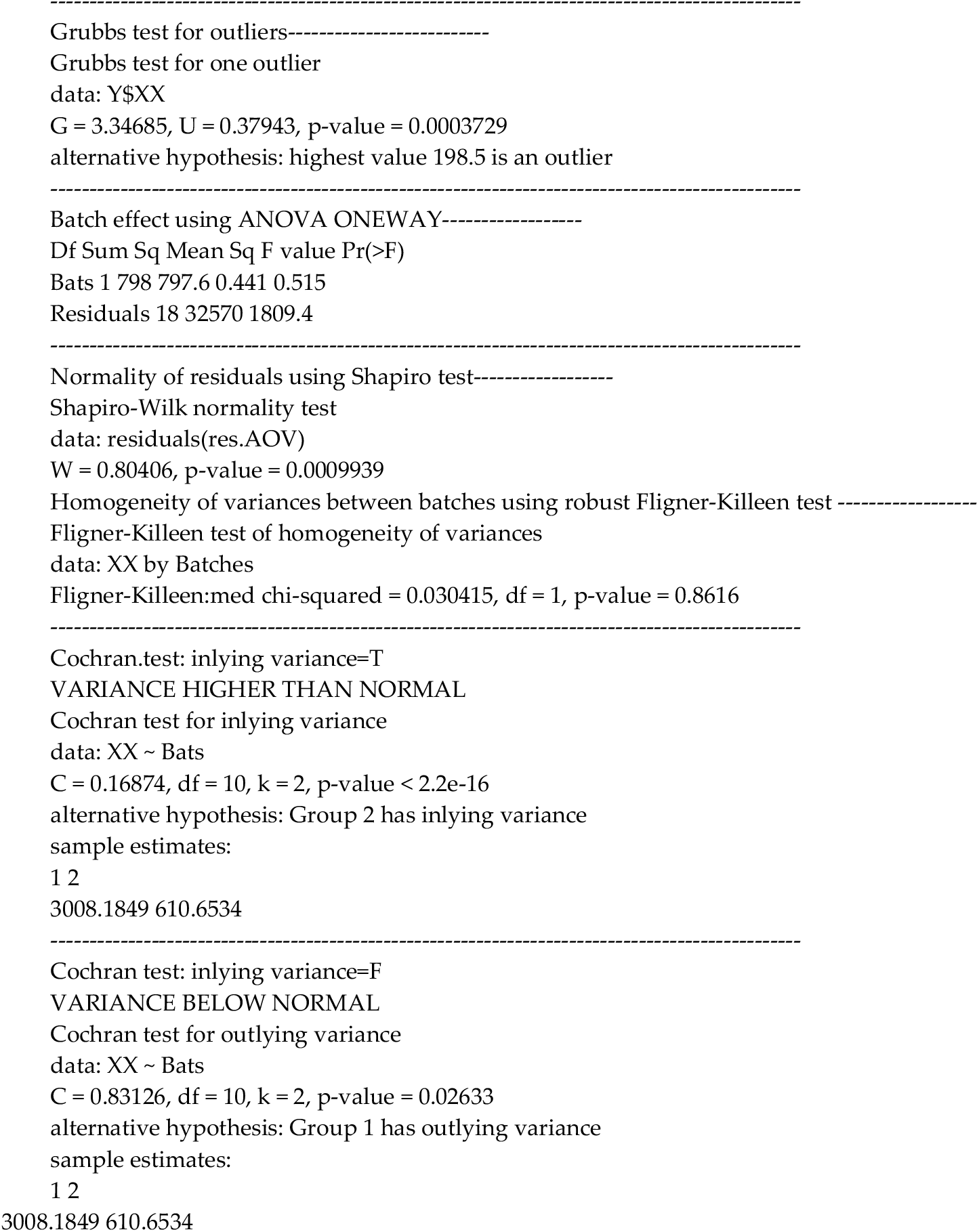

In the results it is possible to appreciate the presence of at least 3 outliers, which correspond to observations 6, 13, 20 (Hampel filter method) or to observations 13, 20 (according to the Box-plot method). Grubbs test values for outliers were significant, indicating that there are outliers within the observations of the response variable. Another relevant result is the evidence of data non-normality (Shapiro test, p<0.05).

This result is complemented with a boxplot that reveals possible outlier values, as can be seen in Figure 7.

**Figure 7:**
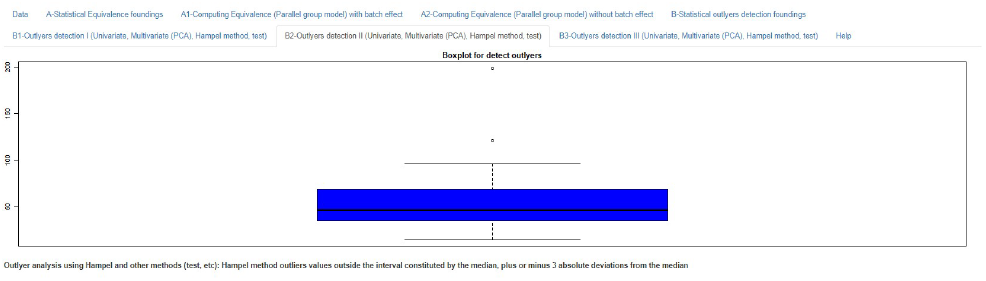
Eq-DIAGNOBATCH: Boxplot of the observations for a parallel group design without a batch effect for the file Eq_Parallel_withoutBatch.csv. Different out-of-range observations can be seen at the top of the box plot (white circles).

### 3.2. Multivariate outlier analysis

In multivariate analysis of outliers, which is advisable when there is more than one response variable in the data, the procedures are more complex. They have been previously cited in the introductory section and are referenced in Appendix 2. This analysis is based on the reduction of the dimension using PCA followed by analysis of the coordinates of the different points with methods such as the Hampel method and the Grubbs test for outliers.

This type of outlier analysis is illustrated by the file Analysis_outlyers_multiv.csv, which contains three response variables (Var1, Var2, Var3). This is an example of equivalence of a pharmacological treatment with different batches that have problems of variability between and within the batches, which must be detected prior to the equivalence analysis in order to explain the equivalence results.

The main result is presented in Figure 8. The diagram represents the percentiles obtained by the Hampel method after converting the variables in two components using PCA and a list of the possible outliers.

**Figure 8.**
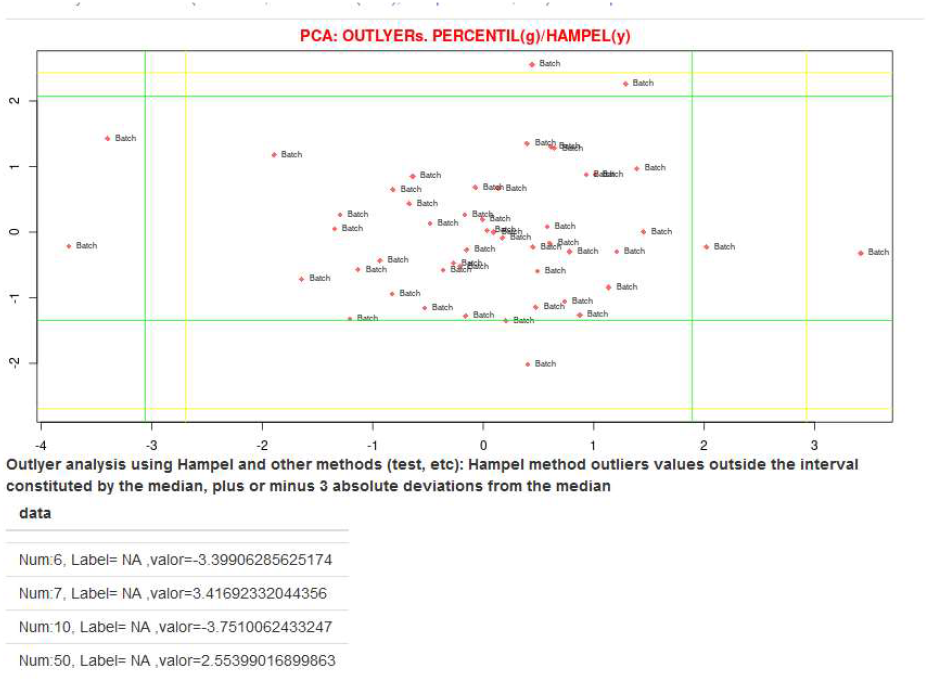
Eq-DIAGNOBATCH: Outlier multivariate analysis using the example Analysis_outlyers_multiv.csv, which contains 3 variables and multiple batches and only one treatment is considered. The percentile diagram obtained by the Hampel method is represented by yellow lines, which correspond to the 3 median limits.

The list of the statistical results where the outliers are detected using different methods (Hampel method and Grubbs test) are also presented by the application:

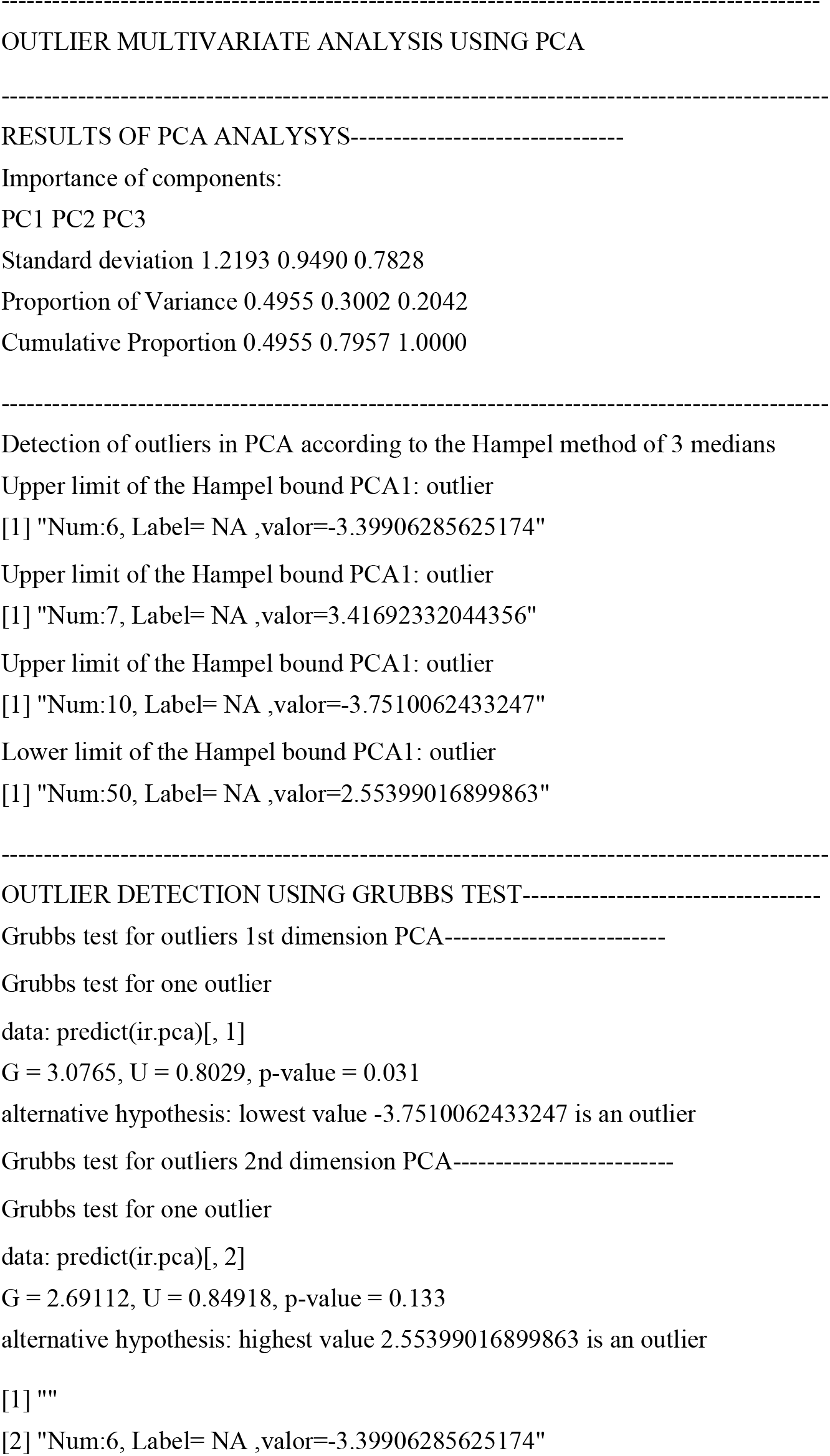
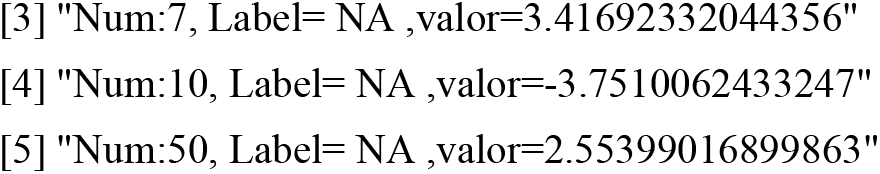

In these results, it is possible to appreciate the presence at least 4 outliers, which correspond to the observations 6, 7, 10, 50 (Hampel filter method) or to observations 6, 7, 10, 50, 20 (according to the Grubbs test). The Grubbs test values for outliers are significant (p=0.031, for the first PCA dimension), indicating there are outliers within the observations of the response variable.

## Conclusions

The Eq-DIAGNOBATCH application presented here complements a previous theoretical article about establishing the equivalence of topical products in a parallel linear model, with or without a batch effect. The application provides a very simple way of performing the equivalence test for this type of situation, efficiently obtaining estimates of the parameters of interest (equivalence intervals, geometric mean ratio, and variance components), and allowing calculations to be carried out, either assuming a batch effect or not. Real-life situations have been presented in which equivalence has been declared and others in which this was not possible due to the existence of great variability. The equivalence limits can be easily changed through an option in the application, which also examines the statistical implications of the presence of outliers in the sample, from both a univariate and multivariate point of view. The former is based on the Hampel method and Grubbs test, while the latter is based on dimension reduction using PCA and later analyzing the coordinates of the different points with methods such as the Hampel approach and Grubbs test. The operation and usability of the application are demonstrated in different examples, taken from theoretical literature or real-life industry case studies. The results seem to confirm the facility of the application to detect outliers, essential when carrying out an equivalence analysis.

## Data Availability

The application is free to use and can be accessed at the following link:
https://amonleongbiost3.shinyapps.io/EqDIAGNOBATCH/. The different usage dataframes can be
found in https://github.com/amonleong/EqDIAGNOBATCH.

## Appendix. Summary of statistical methods used in App Eq-DIAGNOBATCH

This summarizes and complements the statistical theory presented in Ocaña et al (2020) and clarifies the statistical methods used in the *Eq-DIAGNOBATCH* application.

## Appendix A. Experimental design for establishing equivalence in a study of parallel groups and treatment effect (equivalence method for two populations)

### A.1. Experimental design in the case of equivalence (parallel groups)

The most appropriate experimental design describing the process of acquiring data is a one-factor design, with the fixed factor “form”, with *a* = 2 levels: “Test” and “Reference” (*T* and *R* from now on).

The main objective of a test vs. reference equivalence study is to study the “form” factor. The fixed levels to consider are precisely *T* and *R*.

The linear model associated with this design may be written as in Equation (1):

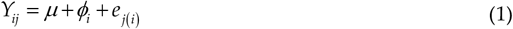

where *Y*_*ij*_ stands for the observed response (e.g., quality measures like viscosity, the AUC measuring spreadability, or a PCA summary of the rheology variables, as discussed in Ocaña et al. (2020)); *μ* stands for the constant or global model mean; the constant *ϕ*_*i*_ corresponds to the fixed effect of the i-th formulation, *i* = *T, R*; and *e*_*j*(*i*)_stands for the residual associated with the j-th replicate observation of the i-th formulation. As is usual, we assume that all *e*_*j*(*i*)_ are mutually independent, with a mean of zero. The common residual variance will be designated as 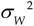. Model 1 is simple and as it does not have more than two groups, with a single fixed factor (treatment) of two levels (T, R), the estimation of the parameters can be simplified.

The mean response for each formulation, *µ*_*T*_ = *µ* + *ϕ*_*T*_ and *µ*_*R*_ = *µ* + *ϕ*_*R*_, may be unbiasedly estimated by the formulae in Equations (2) and (3):

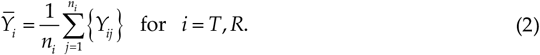

with the overall variance:

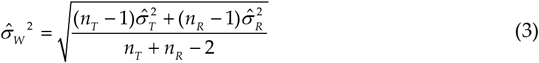

where *n*_*T*_ , *n*_*T*_ are the sample size groups for R and T, and 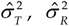 are the group variances for R and T.

If *μ*_*R*_ and *μ*_*R*_ are the population means for the T and R formulations, respectively, and [Δ_1_ ; Δ_2_ ] is the absolute equivalence interval (e.g. [0.8;1.20]), the additive hypothesis for the equivalence test can be formulated as:

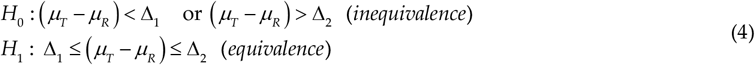

which becomes additive after a log transformation:

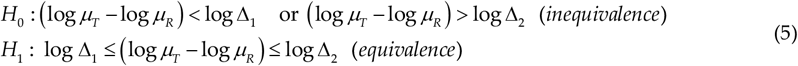

The hyphotesis in (1) can be tested using the two one-sided test (TOST) from Schuirman (1987), where (1) is divided in two hyphotheses, with the first one-sided test:

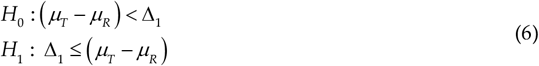

and second one-sided test:

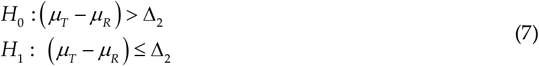

It is possible to declare equivalence when the two tests (6, 7) reject *H*_0_ . The decision rules for the two one-sided test procedure are based on a t-test. In the first one-sided test (3), *H*_0_ is rejected when:

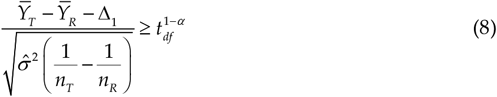

Where 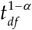 is a T statistic with *n*_*T*_ + *n*_*R*_ − 2 df (df = degrees of freedom) and *α* is the significance level. For the second one-sided test (4), *H*_0_ is rejected when:

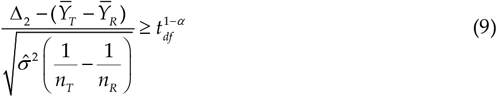

with same interpretation as in formula (5).

In practice, an operationally equivalent procedure is to build a 90% confidence interval (1 − 2*α*) with *α* =0.05 for (*μ* _*T*_ − *μ*_*R*_):

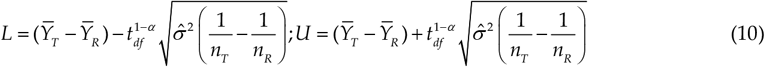

where L is its lower confidence bound and U is its upper confidence bound and 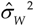 is defined as in (3). Equivalence is declared if this interval lies fully within the equivalence limits, i.e., if L and U are within [Δ_1_; Δ_2_] . We then conclude that the two formulations are equivalent. In general, these values are determined by the regulations or some value based on experience, as is the case of the equivalence of pharmaceutical products for the comparison between T and R.

### A.2. Testing for Quality Equivalence. Ratio of Means Confidence Intervals

Direct Confidence Interval for the Ratio

The EMA (EMA, 2018) draft guideline establishes that the parameter of interest to prove quality equivalence is (*µ*_*T*_ − *µ*_*R*_) / *µ*_*R*_ and suggests [Δ = −0.1; Δ = +0.1]. As a hypothesis testing problem, to prove equivalence corresponds to rejecting a null hypothesis of non-equivalence, Equation (11):

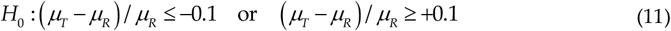

for an alternative of equivalence, Equation (12):

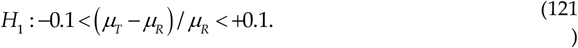

Provided that (*µ*_*T*_ − *µ*_*R*_) / *µ*_*R*_ = *µ*_*T*_ / *µ*_*R*_ − 1, these hypotheses may be restated as in Equation (13):

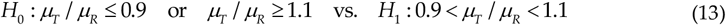

and the problem is reduced to obtaining a 90% confidence interval for the ratio *μ*_*T*_ / *μ*_*R*_ with [Δ = 0.9; Δ = 1.1]

Note that these equivalence limits are symmetric around 1. As they are limits for a ratio of means, it would be more natural to consider asymmetric limits 0.9 to 1/0.9 (=1.1111…), in a similar way to the standard bioequivalence limits 0.8 to 1/0.8 = 1.25. However, for quality equivalence, the draft guidelines state the problem in terms of the difference of means relative to the *R* mean and this naturally leads to the limits stated in (13), which we will consider from now on.

### A.3. Direct Confidence Interval for the Difference of Means

The statement defining the quality equivalence criterion, i.e., (4), may be restated as −0.1*μ*_*R*_ < *μ* _*T*_ − *μ*_*R*_ < +0.1*μ* when [Δ = 0.9; 1.1] .Thus, a simpler but conceptually less correct alternative approach would be to obtain a confidence interval for the difference of means and (by directly substituting the unknown *μ* _*R*_ by its estimate, 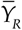) to conclude equivalence if it is fully included in the (random) limits 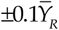. In other words, quality equivalence may be concluded if the condition in Equation (10) is met:

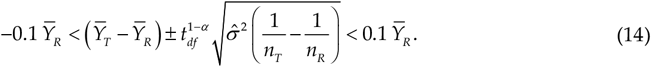

Except for the confidence intervals under consideration, this equivalence criterion is the same as those discussed in the previous subsection. Note that by dividing all terms by 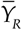 the expression (14) is reduced to requiring that the following confidence interval for the relative difference of means, (*µ*_*T*_ *µ*_*R*_) / *µ*_*R*_, Equation (15),

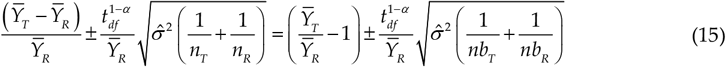

must be fully included between the ±0.1 limits. After some elementary algebraic manipulation, the above criterion is itself reduced to the condition that a confidence interval for the ratio *µ*_*T*_ / *µ*_*R*_ must be fully within the range 0.9 to 1.1 when [Δ_1_ = 0.9; Δ_2_ = 1.1], and 0.8 to 1.25 when [Δ_1_ = 0.8; Δ_2_ = 1.25]

Though commonly used in applications, we will not consider this approach hereafter, but use the most common form based on the logarithmic transformation that gives rise to the Confidence Interval for Log-Transformed Data.

### A.4. Confidence Interval for Log-Transformed Data

Although the draft guidelines clearly state that normality can be assumed for the quality measures, arguments were provided at the EUFEPS open forum [Monleón-Getino et al, 2019] concerning this assumption. If assuming normality on a log scale was more plausible, the ratio of means previously considered should now be viewed as a ratio of geometric means on the original log-normal scale, which becomes a difference of means on the log-transformed scale.

Then the formulation effect, *ϕ* = *ϕ*_*T*_ − *ϕ*_*R*_, which is the mean difference between both forms on a log scale (now all parameters and their estimates refer to these log-transformed data), can be unbiasedly estimated by 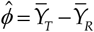 The meaning of all previous symbols remains the same, but is considered or computed for the log-transformed variables.

The procedure may be stated as follows:

1. Log-transform the quality variable under consideration. Now *Y* in model (1) stands for the log-transformed values.
2. With these log-values, compute a 1 – 2*α* confidence interval (typically a 90% interval, with *α* = 0.05) for the form effect, ϕ, on a log scale, Equation (16):
3. 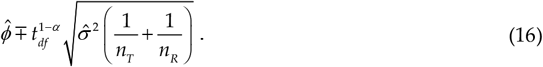
4. Back-transform this confidence interval (*IC_GMR_* to obtain 90% (or more in general 1 – 2*α*) for the geometric means ratio between *T* and *R* on the original scale, Equation (17):

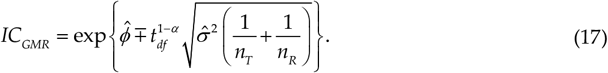
5. Equivalence is declared if *IC*_*GMR*_ is fully included within the equivalence limits 0.9 to 1.1 (or any other limits considered adequate), Equation (18):

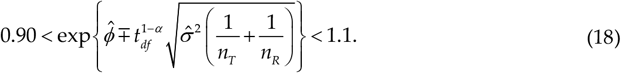

Otherwise, it is not possible to declare equivalence.

Fieller’s confidence interval for the ratio of means was presented in Ocaña et al (2020) as a general way of computing the confidence interval of the ratio of two means *μ*_*A*_ / *μ*_*B*_ . For simplicity, the case included independent normally distributed estimates 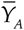 and 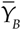 of *μ*_*A*_ and *μ*_*B*_, respectively. Specifically, 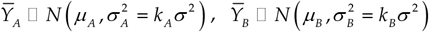 and 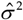 is an estimator of *σ*^2^ with *r* degrees of freedom.

## Appendix

B. Outlier detection methods

The application can detect outliers in two ways: a) in a multivariate approach, using all the response variables (Y), previously performing a PCA, and working only with the first two components (Ocaña et al., 2020) and b) in a univariate approach, using only the response variable required in the analysis. The multivariate analysis is performed automatically when a file with more than one quantitative response variable is loaded. If there is only one variable and it is indicated in the variables option, a univariate analysis of outliers is performed. The application uses common methods to detect outliers (Soetewey, 2020), such as boxplots, Hampel’s filter (Kuppusamy & Kaliyaperumal, 2013; Soetewey, 2020) and statistical tests such as Grubbs test (Grubbs, 1969).

In the multivariate analysis (see Ocaña et al., 2020) the Hampel filter (Kuppusamy & Kaliyaperumal, 2013; Soetewey, 2020) and Grubbs test (Grubbs, 1969) are used to detect outliers in the case of PC1 and PC2. Added to the unidimensional analysis is the analysis of outliers based on the interquartile distances for boxplots. The univariate analysis is also completed by using the ANOVA model to detect a batch effect (random factor), complemented by the robust Fligner-Killeen test **(Conover et al, 1981)** for the homogeneity of variances between batches and the Shapiro test (Shapiro and Wilk, 1965) for the normality of residuals.

## Footnotes

1 R = reference product, T = test product

## Notes

### Competing Interest Statement

The authors have declared no competing interest.

### Funding Statement

This study did not receive any funding

